# PRedicting the EVolution of SubjectIvE Cognitive Decline to Alzheimer’s Disease With machine learning: the PREVIEW study protocol

**DOI:** 10.1101/2023.04.15.23288619

**Authors:** Salvatore Mazzeo, Michael Lassi, Sonia Padiglioni, Alberto Arturo Vergani, Valentina Moschini, Maenia Scarpino, Giulia Giacomucci, Rachele Burali, Carmen Morinelli, Carlo Fabbiani, Giulia Galdo, Silvia Bagnoli, Filippo Emiliani, Assunta Ingannato, Benedetta Nacmias, Sandro Sorbi, Antonello Grippo, Alberto Mazzoni, Valentina Bessi

**Author notes:** corresponding author: Valentina Bessi, Department of Neuroscience, Psychology, Drug Research and Child Health, University of Florence, Azienda Ospedaliero-Universitaria Careggi. Largo Brambilla, 3, 50134, Florence, Italy.

## Abstract

**Background and aims:** Subjective Cognitive Decline (SCD) is a condition in which individual complain of cognitive decline with normal performances on neuropsychological evaluation. Many studies demonstrated a higher prevalence of Alzheimer’s pathology in patients diagnosed with SCD as compared to the general population. Consequently, SCD was suggested as an early symptomatic phase of Alzheimer’s disease (AD). We will describe the study protocol of a prospective cohort study (PREVIEW) that aim to identify features and tools to accurately detect SCD patients who will progress to AD.

**Methods:** We will include patients self-referred to our memory clinic and diagnosed with SCD. Participants will undergo: clinical, neurologic and neuropsychological examination, estimation of cognitive reserve and depression, evaluation of personality traits, *APOE* and *BDNF* genotyping, electroencephalography and event-related potential recording, lumbar puncture for measurement of Aβ_42_, t-tau, and p-tau concentration and Aβ_42_/Aβ_40_ ratio. Recruited patients will have follow-up neuropsychological examination every two years. Collected data will be used to train a machine learning algorithm to define the risk of progression from SCD to MCI and AD.

**Discussion:** There is an urgent need to select cost-effective and easily accessible tools to identify patients at the earliest stages of the disease. Previous studies identified demographic, cognitive, genetic, neurophysiological and brain structure features to stratify SCD patients according to the risk of progression to objective cognitive decline. Nevertheless, only a few studies considered all these features together and applied machine learning approaches on SCD patients.

**Conclusions:** the PREVIEW study aim to identify new cost-effective disease biomarkers (e.g., EEG-derived biomarkers) and define automated algorithm to detect patients at risk for AD in a very early stage of the disease.

## 1. Introduction

Research and clinical practice on Alzheimer’s disease (AD) is at a turning point. As disease modifying therapies (DMTs) for Alzheimer’s disease are becoming available (1), neurologists, researchers and health services will be faced with the predictably increasing demands of diagnostic assessment of patients with cognitive disorders. Moreover, it is a shared understanding that DMT should be administered at the earliest stages of the disease, to stop the pathologic process before neurodegeneration starts (2). For this reason, international research is focusing on prodromal and preclinical phases of AD. Subjective cognitive decline (SCD) was defined as a self-experienced persistent decline in cognitive capacity in comparison with the previously normal status, during which the subject has normal age-, sex-, and education-adjusted performance on standardized cognitive tests (3). SCD was associated with neuroradiological features suggestive of AD (4), amyloid deposition (5,6) and higher risk of progression to Mild Cognitive Impairment (MCI) or dementia as individuals without SCD (7). Based on this evidence, the National Institute of Aging-Alzheimer’s Association (NIA-AA) included SCD as a first manifestation of the symptomatic stages of AD (8), preceding MCI (9). Hence, SCD might represent a target population for DMT to preserve cognitive function and psychological well-being (10). On the other hand, SCD constitutes a heterogeneous group with several possible trajectories (11) and many potential underlying causes including: normal aging, personality traits, other psychiatric, neurologic or medical disorders, substance use, and medications (12). Therefore, it is crucial to identify features and tools to accurately detect prodromal AD among patients with SCD.

This paper describes the protocol of the PREVIEW (PRedicting the EVolution of SubjectIvE Cognitive Decline to Alzheimer’s Disease With machine learning) study that will prospectively investigate baseline predictors and biomarkers of Alzheimer’s pathology and progression to MCI and dementia in a large cohort of patients with SCD.

In recent years, The Regional Reference Centre for Alzheimer’s Disease and Cognitive Disorders of Careggi Hospital (Florence, Italy) analyzed a large set of neuropsychological, personality and lifestyles data from patients with SCD collected in about 25 years, identifying demographic (13,14), cognitive (15,16), personality (15) and genetic (13,17–22) features that increase the risk of progression from SCD to MCI or AD.

In this study we will integrate our previous findings with data from non-invasive techniques, namely electroencephalography (EEG) and event-related potentials (ERP) recording. These techniques reliably measure the neural circuits associated with cognitive processes and may provide sensitive metrics for early diagnosis of cognitive impairment (23). We will apply machine learning approaches, an emerging and promising tool that showed great potential in diagnosis and classification of neurodegenerative diseases and other medical conditions (24–26). We aim to:

i. integrate a multimodal set of data from SCD patients including clinical data, neuropsychological assessment, personality traits, cognitive reserve, genetic factors and features from EEG and ERP recordings;
ii. assess the accuracy of these data in predicting conversion from SCD to MCI and AD through machine learning tools;
iii. define a management protocol for SCD to be applied in memory clinic settings. The PREVIEW study was registered on ClinicalTrials.gov (registration number: NCT05569083).

## 2. Methods and Analysis

### 2.1. Study Design and Participants

This is a longitudinal single center observational cohort study. We will consider consecutive spontaneous patients, who self-referred to the Centre for AD and Adult Cognitive Disorders of Careggi Hospital in Florence, classified as SCD according to SCD-I criteria (27).

We will recruit patients who meet the following criteria:

a. complaining of cognitive decline with a duration of ≥ 6 months;
b. Mini Mental State Examination (MMSE) score greater than 24, corrected for age and education;
c. normal functioning on the Activities of Daily Living (ADL) and the Instrumental Activities of Daily Living (IADL) scales (28);
d. unsatisfied criteria for MCI and AD according to NIA-AA criteria (29,30);
e. give their written informed consent.

Exclusion criteria are:

a. history of head injury, current neurological and/or systemic disease, symptoms of psychosis, major depression, alcoholism or other substance abuse;
b. the complete data loss of patients during follow-up.

All recruited patients will undergo at baseline: comprehensive familial and clinical history, extensive neuropsychological assessment, estimation of premorbid intelligence, assessment of depression, personality and leisure activities evaluation, *APOE* and *BDNF* genotype analysis, EEG and ERP recording. CSF analysis will be performed in patients who will give additional informed consent for lumbar puncture.

Patients will undergo neuropsychological evaluation every two years. Progression to MCI and to AD dementia will be defined according to NIA-AA criteria (29,31). Patients who will progress to dementia will be addressed to diagnostic and therapeutic work-out adopted in our center (**Fig.1**).

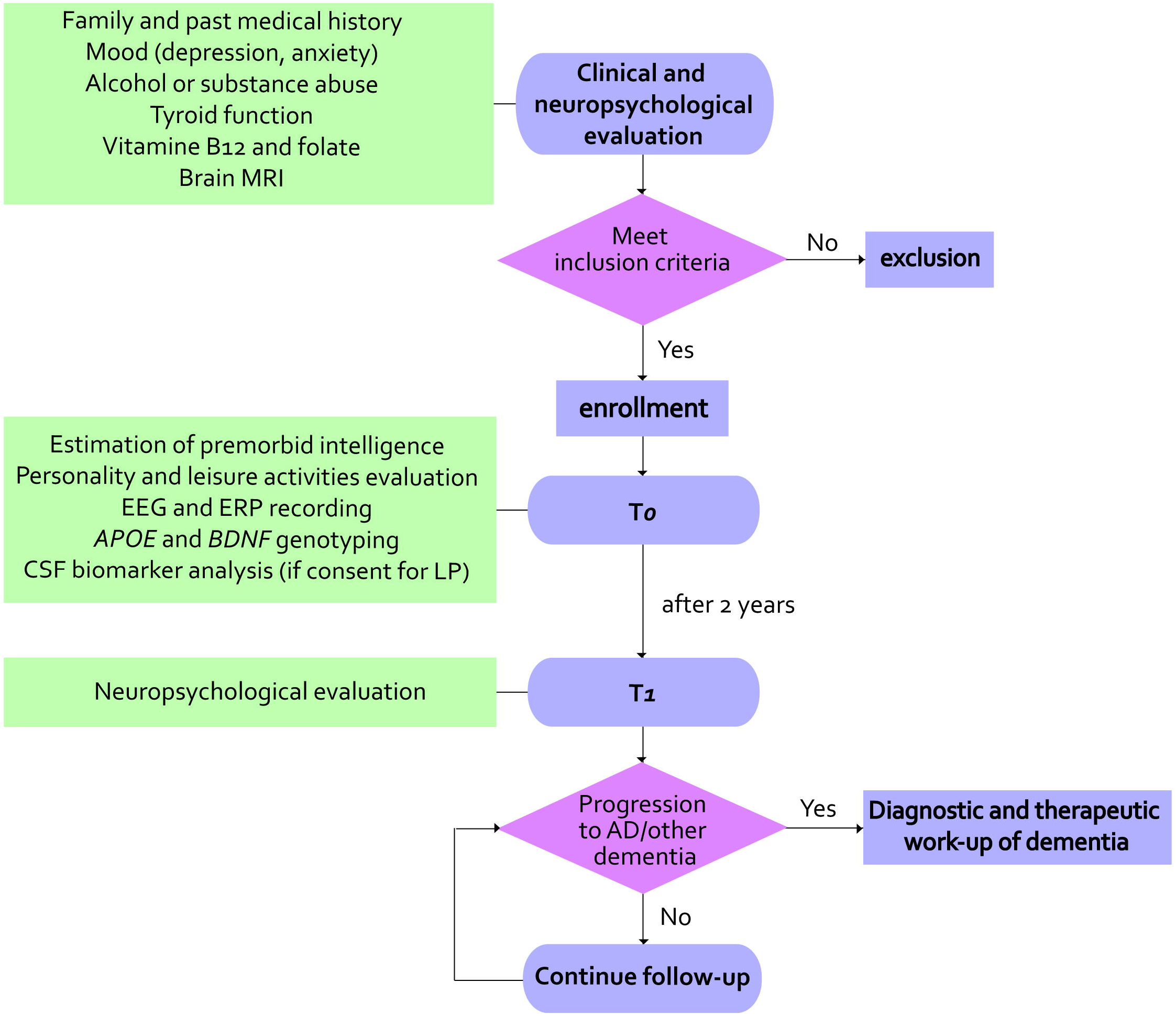

Two samples of i) age-matched healthy controls (without cognitive concern) and ii) MCI patients will undergo EEG and ERP recording for the purpose of cross-sectional comparison with SCD group at baseline.

### 2.2. Neuropysychological evaluation, assessment of depression and estimation of premorbid intelligence

Extensive neuropsychological examination will include: global measurements (MMSE)(32), tasks exploring verbal and spatial short-working and long-term memory (Digit and Visuo-spatial Span forward and backward (33), Rey Auditory Verbal Learning Test – RAVLT (34), Short Story Immediate and Delayed Recall (35), Rey-Osterrieth complex figure recall (36)), attention (Trail Making Test A (37), attentional matrices (38), Multiple Features Targets Cancellation – MFTC (39)), language (Category Fluency Task (40), Phonemic Fluency Task (34) and Italian language battery: Screening for Aphasia NeuroDegeneration – SAND(41)), constructional praxis (Copying Drawings (34), Rey-Osterrieth complex figure copy (36), Clock test (42)) and executive function (Trail Making Test B (37), Stroop Test (43), Frontal Assessment Battery(44)). The subjective perception of memory impairment will be investigated using the Memory Assessment Clinics-Questionnaire – MAC-Q (45). Premorbid intelligence will be estimated using short intelligence test – TIB (46), that has been constructed as an Italian equivalent of the National Adult Reading Test – NART (47). The presence and severity of depressive symptoms will be evaluated by means of the 22-item Hamilton Depression Rating Scale (HDRS) (48). Katz Index of Independence in Activities of Daily Living (ADL) (49) scale will be used to assess functional capacities at baseline and at follow-up.

### 2.3. Personality Traits and Leisure Activities

We will use the Big Five Factors Questionnaire (BFFQ) (50) to assess personality traits. Participants will be asked to fill out a questionnaire that measures the five factors of: 1) emotional stability, 2) energy, 3) conscientiousness, 4) agreeableness, and 5) openness to culture and experience. The inventory follows a widely accepted five-traits personality model (50,51). For the 24 items of each factor, subjects will rate their level of agreement on a five-point scale ranging from strongly agree to strongly disagree. Item scores will be computed for each factor to yield a summary measure of the trait with higher values representing a greater degree of the explored dimension.

At baseline, subjects will be interviewed regarding participation, when they were 30-40 years old, in nine Intellectual Activities, seven Social Activities and seven Physical Activities (modified from Yarnold PR et al. (52)). The frequency of participation will be reported for each activity on a Likert scale ranging from 0 to 5, where 0 refers to never, 1 to occasionally, 2 to monthly, 3 to once a week, 4 to several days per week and 5 to daily.

### 2.4. EEG and ERP recording

Resting-state EEG data will be collected at the Neurophysiological Laboratory of IRCCS Don Gnocchi (Florence, Italy) using the 64-channels Galileo-NT system (E.B. Neuro S.p.a.). The EEG will be recorded continuously from 64 electrodes using an EEG Prewired Headcups. Electrodes were positioned according to the 10–10 international system. (AF7, AF3, Fp1, Fp2, Af4, Af8, F7, F5, F3, F1, F2, F4, F6, F8, FT7, FC5, FC3, FC1,FC2, FC4, FC6, FT8, T3, C5, C3, C1, C2, C4, C6, T4 TP7, CP5, CP3, CP1, CP2, CP4, CP6, TP8, T5, P5, P3, P1, P2, P4, P6, T6, Fpz, PO7, PO3, O1, O2, PO4, PO8, Oz, AFz, Fz, FCz, Cz, CPz, Pz, and POz).The ground electrode will be placed in front of Fz. Horizontal eye movements will be detected by electrooculogram (EOG). Data will be digitized at a sampling rate of 512 Hz and analogue-digital precision will be 16 bits. The recording will be referenced to the common average of all electrodes, excluding Fp1 and Fp2. Re-referencing will be done prior to the EEG artifact detection and analysis. Electrode-skin impedance will be set below 5 kilo-ohms. Subjects will be sat in a reclined chair in a comfortable position. Resting EEG recording begin with a 10-minute eyes-closed registration followed by an alternance of 3 minutes eyes-open and 3 minutes eyes closed, repeated twice. First phase of recording was designed to be longer in order to obtain a reliable resting state recording at eyes closed. Only the eyes-closed portions of the signal will be used for subsequent analyses.

ERP acquisition will be performed with the same the same EEG system used for EEG data acquisition. The participants will be administered an ERP test battery with concurrently recorded EEG consisting of a 3-choice vigilance task (3CVT) and standard image recognition task (SIR). The first is designed to evaluate sustained visual attention and the second designed to evaluate attention, encoding, and image recognition memory. In particular, SIR images were defined as stimuli to distinguish working memory from semantic memory loss and extend previous results of image recognition ERP effects.

In order to remove electrophysiological and non-electrophysiological artifacts from the raw signals, we used a custom preprocessing pipeline written in MATLAB with the use of the EEGLAB toolbox functions (53). The pipeline consisted of two main steps: the PREP pipeline (54), followed by independent component analysis (ICA) to remove artifactual components (55). Moreover, to generate the ERP epochs, time windows from -300 ms to + 1000 ms will be created for each EEG recording channel, with the stimulus presentation centered at 0 ms and the average of the trials related to each epoch will be calculated.

### 2.5. AD biomarker measurement and genetic analysis of *APOE* and *BDNF* genes

Blood and CSF samples will be immediately centrifuged, stored at -80 °C and analyzed at the Laboratory of Neurogenetics of Careggi University Hospital. Aβ_42_, Aβ_42_/Aβ_40_ ratio, t-tau, and p-tau will be measured using a chemiluminescent enzyme immunoassay (CLEIA) analyzer LUMIPULSE G600 (Fujirebio, Tokyo, Japan). Cut-off values for CSF will be determined following Fujirebio guidelines (Diagnostic sensitivity and specificity using clinical diagnosis and follow-up golden standard, November 19^th^, 2018): Aβ_42_ > 670 pg/ml, Aβ_42_/Aβ_40_ ratio > 0.062, t-tau < 400 pg/ml and p-tau < 60 pg/ml. The three SNPs (rs429358, rs7412 and rs6265 on *APOE* and *BDNF* genes respectively) will be analyzed by the polymerase chain reaction (PCR) on genomic DNA and with the analysis of melting curves (HRMA) using the Rotor-Gene 6,000 (Rotor-Gene, Corbett Research, Mortlake, Australia).

### 2.6. Data Collection and Management

Data collection will be carried anonymously on REDCap, an online-based software for the design of databases. Data will be collected in a pseudo-anonymized way, attributing a record ID to each patient on the electronic database and saving the correspondences between names and identification codes on a separate document.

### 2.7. EEG pre-processing

The first step of preprocessing will rely on the PREP pipeline, which performs several preprocessing steps automatically, allowing to obtain robust average re-referenced signals. At first, PREP high-pass filter signals of all channels, by means of a Hamming windowed FIR filter (using EEGLAB’s *pop_eegfiltnew* function) with a 1 Hz cut-off frequency. Line noise at 50 Hz and its harmonics were removed by using the CleanLine EEGLAB plugin. Noisy channels, i.e., those channels having abnormal and/or uncorrelated activity compared to others will be removed by using PREP noisy channel subroutine, which performs the bad channel selection by combining an ensemble of methods: the deviation criterion, the correlation criterion, the noisiness criterion, and the predictability criterion. Remaining channels activity will be used to estimate a robust average reference, based on robust statistics such as the median and interquartile range. Finally, removed channels will be interpolated, by means of spherical interpolation. The obtained re-referenced and filtered signals will be subjected to the second preprocessing step.

Independent components will be extracted by using the Infomax ICA algorithm (57), as implemented in *binica* EEGLAB routine. A semi-automated procedure will be then used to distinguish between brain-related components and artifactual ones. We used ICLabel (58) to classify automatically independent components into brain or artifactual components (line noise, muscle, eye, channel noise, heart, “other”) based on a neural network trained on crowd-sourced data. ICLabel returns the probability of each component to belong to one of the above-mentioned classes. We will then use DIPFIT to perform a single dipole fitting of the independent component map onto a template brain (MNI-152 atlas). Given that brain components should be dipolar (59), a high residual variance of the fitted dipole should indicate a low probability of the component being brain-related. Hence, components labeled by ICLabel as “brain” with a confidence higher than a threshold (75%) and fitting dipole residual variance lower than another threshold (20%) will be retained in the final signals. Noise components with high confidence and high dipole residual variance will be instead automatically removed from the ICs list. All the remaining components will be inspected visually and flagged either as brain or non-brain depending on their power spectra profiles and time-courses. Channel-level signals will be reconstructed from the reduced IC space including only brain-related sources. Finally, we will perform a visual inspection of the cleaned signals, to remove possible remaining artifacts (e.g., temporally localized muscle activity not removed by the ICA procedure).

### 2.8. EEG Statistical Analysis

At first, we will compute the power spectral density (PSD) of the signal in each of the recorded channels, applying the Welch’s method on continuous windows of EEG signals, using Hanning windows with no overlap. We will divide the spectrum in four canonical frequency bands, namely: delta (1-4 Hz), theta (4-8Hz), alpha (8-13 Hz) and beta (13-30 Hz). We will divide the scalp in six regions of interest (ROIs)(60): frontal right (Fp2, AF4, AF8, F2, F4, F6, F8) frontal left (Fp1, AF3, AF7, F1, F3, F5, F7), central right (FC2, FC4, FC6, FT8, C2, C4, C6, T4, CP2, CP4, CP6), central left (FC1,FC3, FC5, FT7, C1, C3, C5, T3, CP1, CP3, CP5), occipital right (P2, P4, P6, T6, PO8, PO4, O2) and occipital left (P1, P3, P5, T5, PO7, PO3, O1). ROI power will be computed as the average relative power from channels belonging to each ROI.

We will extract several network metrics from the weighted undirected adjacency matrices. First, we will extract the average strength of the connectivity among pairs of ROIs, i.e., the mean weight of non-zero connections. Then, the weighted clustering coefficient (C) and weighted characteristic path length (L) will be computed as previously described (61). Finally, we will compute the small-world coefficient

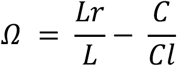

Where L and C are the previously computed weighted clustering coefficient and weighted characteristic path length, whereas Lr is the weighted characteristic path length of an equivalent random network and Cl is the weighted clustering coefficient of an equivalent lattice network.

The last statistical analysis will consist in extracting microstate maps for each subject individually. We will extract a first set of common microstates from the entirety of subjects (independent of conditions), by using the modified k-means algorithm (62). To reduce the computation time, 1000 global field power (GFP) peaks will be randomly selected as input topographies to the clustering algorithm, as these maps should have the highest signal-to-noise ratio. GFP is defined (63) as the standard deviation of all electrodes’ potentials at a given time. Topographies having GFP higher than two standard deviations from the mean will be then discarded from the selection process, as to avoid introducing noisy maps in the clustering algorithm. We will run the modified k-means (20 repetitions, 1000 max iterations) for each number of clusters from two to fifteen. For each iteration of the process, the found microstate maps were back-fitted to the entire preprocessed EEG signal, by assigning each topography from each time-point to the one minimizing the global map dissimilarity (GMD), namely the GFP of the difference between the two maps. The global explained variance will be computed for each topography set. Finally, we will select the number of microstates for which the local improvement in the global explained variance metric will be less than 5%. Once we will determine the globally optimal number of maps, we will fix this number for all subsequent analyses, as to allow for easier comparison between groups. The microstate extraction will be performed using the EEGLAB Microstate plugin, while the back-fitting with the Microstate EEGLAB toolbox (64).

Spectral features, network metrics and microstate features (duration, transition probability) will be compared across conditions and will lay the ground for the machine learning analysis.

### 2.9. Machine learning classification of patient conditions

Machine learning analysis will proceed as follows. We will define a set of multi-modal features including all the features extracted from EEG (see previous subsection), neuropsychological tests, personality traits and genetic variants. We will define a common metric based on the dispersion of each single dimension of the profile and then we will train a machine learning algorithm to associate each profile “vector” to the associated evolution of the disease after a fixed time (at first the possible categories will be only SCD, MCI, AD with no further sublevels, but this can be improved in further rounds). First, we will perform this classification with standard machine learning procedures as support vector machine (SVM, using binary decision trees) and k-nearest neighbors (kNN). In a separate set of analysis, we will follow a deep learning approach, using the same subject profiles to train a multi-level feedforward artificial neural network (ANN) predicting the condition of the patient after a fixed time. The networks will be trained with standard gradient descent and backpropagation techniques and by means of dropout and batch normalization procedures we will obtain robust automatic classification results. Comparing Machine learning and ANN classification performance we will decide the most convenient approach for the task. Moreover, we will apply standard dimensionality reduction techniques to extract the most salient features and repeat the procedure described above to assess whether it is possible to achieve the same results with a subset of the screenings. Once the classification at the current state will be performed, we will exploit longitudinal data to develop an algorithm able to predict the future evolution of cognitive impairment. For instance, an algorithm trained to discriminate between healthy subjects and MCI patients could be applied to SCD patients and evaluate the risk of progression of the patient based on the distance relative to the two clusters.

## 3. Discussion

On June 7, 2021, the Food and Drug Administration (FDA) granted provisional approval of aducanumab, an anti-amyloid monoclonal antibody, for the treatment of patients with MCI due to AD and mild AD dementia (65). Two more monoclonal antibodies, donanemab (66) and lecanemab (67), are under review by the FDA and additional new treatments for AD may become available in the foreseeable future (1). These treatments are not without risk, with amyloid-related imaging abnormality (ARIA) as the most common adverse effect (68). In this scenario, there is an urgent need to select cost-effective and easily accessible tools to identify patients at the earliest stages of the disease trying to minimize, as far as possible, the inclusion of patients who will not progress to AD. The currently recognized disease biomarkers of AD, such as PET neuroimaging (69–71) or CSF biomarkers (72,73), are expensive, almost invasive and, therefore, not suitable to be applied on large populations. At the same time, a general population screening might lead to an unacceptable amount of false positive results and consequent further costs. In this perspective, patients with subjective cognitive decline (SCD) represent an optimal selected population to be screened for prodromal AD. Many studies explored the demographic, cognitive, genetic, neurophysiological and brain structure features potentially associated with the probability that SCD was due to AD and the risk of progression to objective cognitive decline. Regarding demographic and genetic features, the SCD-Initiative proposed a set of features (SCD-plus) to increase the likelihood that SCD indicates pathological change due to AD (27). Our research group of the Regional Reference Centre for Alzheimer’s Disease and Cognitive Disorders of Careggi Hospital previously showed that combining age and *APOE* genotype allow to stratify SCD patients according to the risk of conversion to AD (13). Other genes have also been identified to be linked with changes in cognition in SCD such as *KIBRA* (17), *BDNF* (19), *CLOCK, PER2* (18,21) and *HTT* (22,74). Other studies focused on the role of neuropsychometric scores: data from a community based study by Hao et al. including 84 healthy controls and 517 subjects with SCD showed that individuals classified as SCD-plus had poorer performances in global cognitive scores and in long-term verbal memory tasks (75). The DELCODE study, an observational longitudinal multicenter study including 209 healthy controls and 240 SCD patients, showed that subjects experiencing SCD performed worse than healthy controls in tests assessing memory, executive functions and language abilities (76). Poptsi et al. showed that tasks assessing working memory components, inhibition and switching and cognitive flexibility were able to differentiate SCD by older adults without SCD (77). However, only a few longitudinal studies assessed the prognostic value of neuropsychological assessment at baseline, often with conflicting results (78–81). In a previous study, we analyzed data from 287 patients with SCD and MCI showing that SCD patients who progressed to MCI and AD obtained significantly lower scores in neuropsychological tests for long-term verbal memory and ecological memory at the baseline evaluation compared to SCD who remained stable (15).

Several studies reported that lifetime experiences, such as years of education (82), occupational attainment and engagement in mentally stimulating leisure activities (15), may protect against dementia. These findings have been interpreted in the context of the cognitive reserve hypothesis, which assume that highly intelligent or educated individuals appear to be able to better cope with the presence of a neurodegenerative pathology, maintaining a normal functional level for a longer time than less educated people (83). Many studies focused on the relationship between SCD and cognitive reserve (84–88). Our group demonstrated that intellectual activities carried out by patients in previous decades and higher premorbid intelligence act as protective factors, reducing the risk of progression from SCD to MCI (15,16).

The interaction between SCD and mood disorders is controversial. Two recent community-based studies on very large populations showed that depressive symptoms increase the risk of progression to objective cognitive decline and dementia in SCD patients (89,90). A meta-analysis by Huang et al. showed that depression was significantly higher in individuals with SCD than in normal individuals but there was no difference between SCD and MCI or between SCD converters and non-converters (91). In a 7-years of follow-up study of more than 13000 cognitively normal individuals over 50-years old, SCD and depression were independently associated with the risk of developing MCI or dementia but the risk was highest when both depression and SCD coexisted at baseline (90).

Studies on personality traits showed conflicting results about the association with SCD. Most studies agree that high conscientiousness and low neuroticism are associated with a reduced risk of incident AD (92,93). On the contrary, we found that emotional stability was significantly higher in SCD who progressed to MCI or AD (94).

EEG was extensively evaluated as a toll for the diagnosis of dementia in past decades (95–97). Nevertheless, previous studies mostly tried to identify quantitative EEG markers of AD as compared to healthy controls. These markers were classified into four main categories: i) spectral markers, ii) connectivity and network (and/or connectivity) metrics, iii) complexity measures and iv) microstates (98). Regarding spectral markers, slowing in oscillations of the EEG activity was observed, with a decrease of higher frequency activity (alpha and beta bands) or increase of low-frequency power (delta and theta bands) in AD and MCI groups compared to healthy controls (99–101). Other studies showed a reduction in complexity of the EEG signal throughout the development of dementia (102–104). Connectivity studies looked for covariation patterns in EEG sensor or source signals: they found that connectivity between brain areas, specifically in the higher frequency bands (for spectral-related connectivity measures) decreased as cognitive impairment progressed (105–108). More recently, ERP was suggested as a potential sensitive and robust biomarker to track disease progression and to evaluate response to therapy (109). ERPs are reflections of summated postsynaptic inhibitory and excitatory membrane potentials primarily generated by cortical pyramidal cells. Characteristic time-locked ERP waveforms are elicited in response to sensory, motor, and cognitive events (110). Early components (50–200 ms while the stimulus is encoded) reflect sensory processing of the characteristics of the stimuli but can be influenced to some extent by arousal and attention (111). Late ERP components (P300, N400, P600, and late positive potential [LPP]) reflect feature evaluation, memory matching, and processing speed (112). Multiple reports suggest that abnormal amplitude and latency of LPP are associated with cognitive decline (113). LPP is believed to reflect memory encoding and retrieval with possible sources located in the parahippocampal gyrus, medial temporal lobe, and posterior cingulate gyrus, commonly affected by AD (114). To the best of our knowledge only few studies described quantitative EEG changes in patients with SCD (115–117) while ERP was not been investigated in this population.

Demographic, personality, cognitive, genetic, EEG and ERP features which we will collect for the PREVIEW study will be used to train a machine learning algorithm. Other studies already adopted this approach to predict dementia in non-demented population (118,119) and positive AD biomarkers in patients with MCI (120,121). Most studies demonstrated that machine learning algorithms are able to classify images from AD, MCI, and healthy participants with very high accuracy levels (122,123). Nevertheless, only few studies focused on predicting progression of cognitive decline (124–127) and we are not aware of studies that applied machine learning in predicting progression from SCD to dementia. This approach will allow us to consider all collected variables at once, allowing reducing redundancies between them and minimizing the costs for an optimal performance. This represents the main strength and the first innovative outcome of our project. The collection of AD CSF biomarkers is a second relevant strength of our study for two main reasons: i) as previous studies reported that SCD patients develop AD in an average time of ten years (7,94), a long follow-up period will be needed to reach an acceptable sample size of patients progressed to AD. Considering CSF biomarkers, we could identify patients at higher risk for AD (according to the ATN system (128)) and classify them as prodromal AD. This classification might be considered as a surrogate target for preliminary cross-sectional analyses; ii) as stated before, SCD may also represent the first clinical manifestation of medical conditions other than AD. CSF biomarkers will allow us to distinguish SCD with Alzheimer’s pathology from SCD due to other causes.

It is to be noted that this study will include patients that are self-referred to our memory clinic, as recommended by previous studies, to reduce the heterogenicity of the sample and to increase the chances of finding subjects with preclinical AD in comparison with community-based studies (129).

Finally, we would like to stress that all the features that we will consider as possible predictors will be derived from non-invasive, relatively inexpensive and easily accessible techniques. This will allow us to define a predictive model that may serve as a cost-effective and globally scalable tool for a first step in a diagnostic pathway before confirmation of AD pathology via more invasive and expensive tests.

Our project has some limitations: i) CSF will not be available for all the patients; ii) neuroimaging techniques will be used only for basal assessment of patients and will not be considered for machine learning analysis; iii) healthy controls will undergo only EEG and ERP recording.

## 4. Conclusion

The upcoming of DMTs will lead to a radical change in management of patients with cognitive decline due to AD. As the effectiveness of these drugs is strongly linked to the disease stage, clinicians should be ideally able to detect individuals at risk of AD before neurodegeneration begins. Individuals experiencing cognitive complaint without objective demonstration of impairment may represent the perfect target population for this purpose. PREVIEW studies will involve SCD patients extensively characterized through clinical, neuropsychological, neurophysiological, and genetic assessment. The identified potential biomarkers will be evaluated by a machine learning approach aiming to design the most reliable model to predict the risk of progression to AD.

## 5. Ethics and Dissemination

All subjects will be recruited in accordance with the Declaration of Helsinki and with the ethical standards of the Committee on Human Experimentation of Careggi University Hospital (Florence, Italy). The study was approved by the local Institutional Review Board (reference 15691oss). All participants in this study will sign an informed consent, agreeing to participate and to share the results deriving from their data.

## Data Availability

Data derived from this study will be available from the corresponding author upon reasonable request.

## Author contributions

SM, SP, SS, SM, AG and VB contributed to conception and design of the study. SM, ML, SP, VM, GGa, CF, organized data collection. SM, SP, VM, GGi, MS, CM, RB, FE, SB, AI, BN, SS, AG, VB will contribute to data acquisition. AL and AAV will perform the statistical analysis. SM wrote the first draft of this manuscript. ML, SP, AM, AG, AAV wrote sections of the manuscript. All authors contributed to manuscript revision, read, and approved the submitted version.

## References

1. Cummings J, Lee G, Nahed P, Kambar Mezn, Zhong K, Fonseca J, et al. Alzheimer’s disease drug development pipeline: 2022. Alzheimer’s & Dementia: Translational Research & Clinical Interventions. 2022;8(1):e12295.

2. Guest FL, Rahmoune H, Guest PC. Early Diagnosis and Targeted Treatment Strategy for Improved Therapeutic Outcomes in Alzheimer’s Disease. In: Guest PC, curatore. Reviews on New Drug Targets in Age-Related Disorders [Internet]. Cham: Springer International Publishing; 2020 [citato 27 novembre 2022]. p. 175–91. (Advances in Experimental Medicine and Biology). Disponibile su: https://doi.org/10.1007/978-3-030-42667-5_8

3. Jessen F, Amariglio RE, van Boxtel M, Breteler M, Ceccaldi M, Chételat G, et al. A conceptual framework for research on subjective cognitive decline in preclinical Alzheimer’s disease. Alzheimers Dement. novembre 2014;10(6):844–52.

4. Perrotin A, Mormino EC, Madison CM, Hayenga AO, Jagust WJ. Subjective cognition and amyloid deposition imaging: a Pittsburgh Compound B positron emission tomography study in normal elderly individuals. Arch Neurol. febbraio 2012;69(2):223–9.

5. Amariglio RE, Becker JA, Carmasin J, Wadsworth LP, Lorius N, Sullivan C, et al. Subjective cognitive complaints and amyloid burden in cognitively normal older individuals. Neuropsychologia. ottobre 2012;50(12):2880–6.

6. Wen C, Bi YL, Hu H, Huang SY, Ma YH, Hu HY, et al. Association of Subjective Cognitive Decline with Cerebrospinal Fluid Biomarkers of Alzheimer’s Disease Pathology in Cognitively Intact Older Adults: The CABLE Study. J Alzheimers Dis. 2022;85(3):1143–51.

7. Mitchell AJ, Beaumont H, Ferguson D, Yadegarfar M, Stubbs B. Risk of dementia and mild cognitive impairment in older people with subjective memory complaints: meta-analysis. Acta Psychiatr Scand. dicembre 2014;130(6):439–51.

8. Jessen F, Amariglio RE, van Boxtel M, Breteler M, Ceccaldi M, Chételat G, et al. A conceptual framework for research on subjective cognitive decline in preclinical Alzheimer’s disease. Alzheimer’s & dementia : the journal of the Alzheimer’s Association. 2014;10(6):844–52.

9. Albert MS, DeKosky ST, Dickson D, Dubois B, Feldman HH, Fox NC, et al. The diagnosis of mild cognitive impairment due to Alzheimer’s disease: recommendations from the National Institute on Aging-Alzheimer’s Association workgroups on diagnostic guidelines for Alzheimer’s disease. Alzheimers Dement. maggio 2011;7(3):270–9.

10. Bhome R, Berry AJ, Huntley JD, Howard RJ. Interventions for subjective cognitive decline: systematic review and meta-analysis. BMJ Open. 1 luglio 2018;8(7):e021610.

11. Jessen F, Amariglio RE, Buckley RF, van der Flier WM, Han Y, Molinuevo JL, et al. The characterisation of subjective cognitive decline. Lancet Neurol. marzo 2020;19(3):271–8.

12. Margolis SA, Kelly DA, Daiello LA, Davis J, Tremont G, Pillemer S, et al. Anticholinergic/Sedative Drug Burden and Subjective Cognitive Decline in Older Adults at Risk of Alzheimer’s Disease. The Journals of Gerontology: Series A [Internet]. Settembre 2020 [citato 18 settembre 2020];(glaa222). Disponibile su: https://doi.org/10.1093/gerona/glaa222

13. Mazzeo S, Padiglioni S, Bagnoli S, Carraro M, Piaceri I, Bracco L, et al. Assessing the effectiveness of subjective cognitive decline plus criteria in predicting the progression to Alzheimer’s disease: an 11-year follow-up study. Eur J Neurol. maggio 2020;27(5):894–9.

14. Giacomucci G, Mazzeo S, Padiglioni S, Bagnoli S, Belloni L, Ferrari C, et al. Gender differences in cognitive reserve: implication for subjective cognitive decline in women. Neurol Sci. 8 ottobre 2021;

15. Bessi V, Mazzeo S, Padiglioni S, Piccini C, Nacmias B, Sorbi S, et al. From Subjective Cognitive Decline to Alzheimer’s Disease: The Predictive Role of Neuropsychological Assessment, Personality Traits, and Cognitive Reserve. A 7-Year Follow-Up Study. J Alzheimers Dis. 16 maggio 2018;

16. Mazzeo S, Padiglioni S, Bagnoli S, Bracco L, Nacmias B, Sorbi S, et al. The dual role of cognitive reserve in subjective cognitive decline and mild cognitive impairment: a 7-year follow-up study. J Neurol. febbraio 2019;266(2):487–97.

17. Mazzeo S, Bessi V, Padiglioni S, Bagnoli S, Bracco L, Sorbi S, et al. KIBRA T allele influences memory performance and progression of cognitive decline: a 7-year follow-up study in subjective cognitive decline and mild cognitive impairment. Neurol Sci. 5 aprile 2019;

18. Mazzeo S, Bessi V, Bagnoli S, Giacomucci G, Balestrini J, Padiglioni S, et al. Dual Effect of PER2 C111G Polymorphism on Cognitive Functions across Progression from Subjective Cognitive Decline to Mild Cognitive Impairment. Diagnostics (Basel). 18 aprile 2021;11(4):718.

19. Bessi V, Mazzeo S, Bagnoli S, Padiglioni S, Carraro M, Piaceri I, et al. The implication of BDNF Val66Met polymorphism in progression from subjective cognitive decline to mild cognitive impairment and Alzheimer’s disease: a 9-year follow-up study. Eur Arch Psychiatry Clin Neurosci. giugno 2020;270(4):471–82.

20. Ingannato A, Bagnoli S, Bessi V, Ferrari C, Mazzeo S, Sorbi S, et al. Intermediate alleles of HTT: A new pathway in longevity. Journal of the Neurological Sciences [Internet]. 15 luglio 2022 [citato 11 ottobre 2022];438. Disponibile su: https://www.jns-journal.com/article/S0022-510X(22)00136-8/fulltext

21. Bessi V, Giacomucci G, Mazzeo S, Bagnoli S, Padiglioni S, Balestrini J, et al. PER2 C111G polymorphism, cognitive reserve and cognition in subjective cognitive decline and mild cognitive impairment. A 10-year follow-up study. Eur J Neurol. 8 settembre 2020;

22. Mazzeo S, Emiliani F, Bagnoli S, Padiglioni S, Conti V, Ingannato A, et al. Huntingtin gene intermediate alleles influence the progression from subjective cognitive decline to mild cognitive impairment: A 14-year follow-up study. Eur J Neurol. giugno 2022;29(6):1600–9.

23. Ferreira D, Jelic V, Cavallin L, Oeksengaard AR, Snaedal J, Høgh P, et al. Electroencephalography Is a Good Complement to Currently Established Dementia Biomarkers. DEM. 2016;42(1–2):80–92.

24. Samper-González J, Burgos N, Bottani S, Fontanella S, Lu P, Marcoux A, et al. Reproducible evaluation of classification methods in Alzheimer’s disease: Framework and application to MRI and PET data. Neuroimage. dicembre 2018;183:504–21.

25. Bansal A, Padappayil RP, Garg C, Singal A, Gupta M, Klein A. Utility of Artificial Intelligence Amidst the COVID 19 Pandemic: A Review. J Med Syst. 2020;44(9):156.

26. Pellegrini E, Ballerini L, Hernandez Mdcv, Chappell FM, González-Castro V, Anblagan D, et al. Machine learning of neuroimaging for assisted diagnosis of cognitive impairment and dementia: A systematic review. Alzheimers Dement (Amst). 2018;10:519–35.

27. Jessen F, Amariglio RE, van Boxtel M, Breteler M, Ceccaldi M, Chételat G, et al. A conceptual framework for research on subjective cognitive decline in preclinical Alzheimer’s disease. Alzheimers Dement. novembre 2014;10(6):844–52.

28. Lawton MP, Brody EM. Assessment of Older People: Self-Maintaining and Instrumental Activities of Daily Living. The Gerontologist. 21 settembre 1969;9(3 Part 1):179–86.

29. McKhann GM, Knopman DS, Chertkow H, Hyman BT, Jack CR, Kawas CH, et al. The diagnosis of dementia due to Alzheimer’s disease: recommendations from the National Institute on Aging-Alzheimer’s Association workgroups on diagnostic guidelines for Alzheimer’s disease. Alzheimers Dement. maggio 2011;7(3):263–9.

30. Albert MS, DeKosky ST, Dickson D, Dubois B, Feldman HH, Fox NC, et al. The diagnosis of mild cognitive impairment due to Alzheimer’s disease: Recommendations from the National Institute on Aging-Alzheimer’s Association workgroups on diagnostic guidelines for Alzheimer’s disease. Alzheimers Dement. maggio 2011;7(3):270–9.

31. Albert MS, DeKosky ST, Dickson D, Dubois B, Feldman HH, Fox NC, et al. The diagnosis of mild cognitive impairment due to Alzheimer’s disease: recommendations from the National Institute on Aging-Alzheimer’s Association workgroups on diagnostic guidelines for Alzheimer’s disease. Alzheimers Dement. maggio 2011;7(3):270–9.

32. Magni E, Binetti G, Bianchetti A, Rozzini R, Trabucchi M. Mini-Mental State Examination: a normative study in Italian elderly population. Eur J Neurol. maggio 1996;3(3):198–202.

33. Monaco M, Costa A, Caltagirone C, Carlesimo GA. Forward and backward span for verbal and visuo-spatial data: standardization and normative data from an Italian adult population. Neurol Sci. maggio 2013;34(5):749–54.

34. Carlesimo GA, Caltagirone C, Gainotti G. The Mental Deterioration Battery: normative data, diagnostic reliability and qualitative analyses of cognitive impairment. The Group for the Standardization of the Mental Deterioration Battery. Eur Neurol. 1996;36(6):378–84.

35. De Renzi E, Faglioni P, Ruggerini C. Prove di memoria verbale di impiego clinico per la diagnosi di amnesia. 1977;

36. Caffarra P, Vezzadini G, Dieci F, Zonato F, Venneri A. Rey-Osterrieth complex figure: normative values in an Italian population sample. Neurol Sci. 1 marzo 2002;22(6):443–7.

37. Giovagnoli AR, Del Pesce M, Mascheroni S, Simoncelli M, Laiacona M, Capitani E. Trail making test: normative values from 287 normal adult controls. Ital J Neurol Sci. agosto 1996;17(4):305–9.

38. Della Sala S, Laiacona M, Spinnler H, Ubezio C. A cancellation test: its reliability in assessing attentional deficits in Alzheimer’s disease. Psychol Med. novembre 1992;22(4):885–901.

39. Marra C, Gainotti G, Scaricamazza E, Piccininni C, Ferraccioli M, Quaranta D. The Multiple Features Target Cancellation (MFTC): an attentional visual conjunction search test. Normative values for the Italian population. Neurol Sci. febbraio 2013;34(2):173–80.

40. Novelli G, Papagno C, Capitani E, Laiacona M. Tre test clinici di ricerca e produzione lessicale. Taratura su sogetti normali. / Three clinical tests to research and rate the lexical performance of normal subjects. Archivio di Psicologia, Neurologia e Psichiatria. 1 gennaio 1970;477–506.

41. Catricalà E, Gobbi E, Battista P, Miozzo A, Polito C, Boschi V, et al. SAND: a Screening for Aphasia in NeuroDegeneration. Development and normative data. Neurological sciences : official journal of the Italian Neurological Society and of the Italian Society of Clinical Neurophysiology. agosto 2017;38(8):1469–83.

42. Shulman KI, Pushkar Gold D, Cohen CA, Zucchero CA. Clock-drawing and dementia in the community: A longitudinal study. International Journal of Geriatric Psychiatry. 1993;8(6):487–96.

43. Caffarra P, Vezzadini G, Dieci F, Zonato F, Venneri A. Una versione abbreviata del test di Stroop: Dati normativi nella popolazione Italiana. Rivista di neurologia. luglio 2002;12(4):111–5.

44. Appollonio I, Leone M, Isella V, Piamarta F, Consoli T, Villa ML, et al. The Frontal Assessment Battery (FAB): normative values in an Italian population sample. Neurol Sci. giugno 2005;26(2):108–16.

45. Crook TH, Feher EP, Larrabee GJ. Assessment of memory complaint in age-associated memory impairment: the MAC-Q. Int Psychogeriatr. 1992;4(2):165–76.

46. Colombo L, Brivio C, Benaglio I, Siri S, Capp SF. Alzheimer patients’ ability to read words with irregular stress. Cortex. dicembre 2000;36(5):703–14.

47. Bright P, Hale E, Gooch VJ, Myhill T, van der Linde I. The National Adult Reading Test: restandardisation against the Wechsler Adult Intelligence Scale-Fourth edition. Neuropsychol Rehabil. settembre 2018;28(6):1019–27.

48. Hamilton M. A rating scale for depression. J Neurol Neurosurg Psychiatr. febbraio 1960;23:56–62.

49. Katz S, Ford AB, Moskowitz RW, Jackson BA, Jaffe MW. Studies of Illness in the Aged: The Index of ADL: A Standardized Measure of Biological and Psychosocial Function. JAMA. Settembre 1963;185(12):914–9.

50. Goldberg LR. The development of markers for the Big-Five factor structure. Psychological Assessment. 1992;4(1):26–42.

51. Costa PT, McCrae RR. The NEO personality inventory: Manual, form S and form R [Internet]. Psychological Assessment Resources; 1985. Disponibile su: http://scholar.google.com/scholar?cluster=9664024015502458954&hl=en&oi=scholarr

52. Yarnold PR, Stille FC, Martin GJ. Cross-sectional psychometric assessment of the Functional Status Questionnaire: use with geriatric versus nongeriatric ambulatory medical patients. Int J Psychiatry Med. 1995;25(4):305–17.

53. Delorme A, Makeig S. EEGLAB: an open source toolbox for analysis of single-trial EEG dynamics including independent component analysis. Journal of Neuroscience Methods. marzo 2004;134(1):9–21.

54. Bigdely-Shamlo N, Mullen T, Kothe C, Su KM, Robbins KA. The PREP pipeline: standardized preprocessing for large-scale EEG analysis. Frontiers in Neuroinformatics. 2015;9(JUNE):1–19.

55. Delorme A, Sejnowski T, Makeig S. Enhanced detection of artifacts in EEG data using higher-order statistics and independent component analysis. Neuroimage. 15 febbraio 2007;34(4):1443–9.

56. Alcolea D, Pegueroles J, Muñoz L, Camacho V, López-Mora D, Fernández-León A, et al. Agreement of amyloid PET and CSF biomarkers for Alzheimer’s disease on Lumipulse. Annals of Clinical and Translational Neurology. 2019;6(9):1815–24.

57. Bell AJ, Sejnowski TJ. An information-maximization approach to blind separation and blind deconvolution. Neural Comput. novembre 1995;7(6):1129–59.

58. Pion-Tonachini L, Kreutz-Delgado K, Makeig S. ICLabel: An automated electroencephalographic independent component classifier, dataset, and website. Neuroimage. settembre 2019;198:181–97.

59. Delorme A, Palmer J, Onton J, Oostenveld R, Makeig S. Independent EEG Sources Are Dipolar. PLOS ONE. 15 febbraio 2012;7(2):e30135.

60. Fanciullacci C, Bertolucci F, Lamola G, Panarese A, Artoni F, Micera S, et al. Delta Power Is Higher and More Symmetrical in Ischemic Stroke Patients with Cortical Involvement. Frontiers in Human Neuroscience. 2017;0:385.

61. Vecchio F, Miraglia F, Iberite F, Lacidogna G, Guglielmi V, Marra C, et al. Sustainable method for Alzheimer dementia prediction in mild cognitive impairment: Electroencephalographic connectivity and graph theory combined with apolipoprotein E. Ann Neurol. agosto 2018;84(2):302–14.

62. Pascual-Marqui RD, Michel CM, Lehmann D. Segmentation of Brain Electrical Activity into Microstates; Model Estimation and Validation. IEEE Transactions on Biomedical Engineering. 1995;42(7):658–65.

63. Murray MM, Brunet D, Michel CM. Topographic ERP analyses: A step-by-step tutorial review [Internet]. Brain Topogr; 2008. Disponibile su: https://pubmed.ncbi.nlm.nih.gov/18347966/

64. Poulsen AT, Pedroni A, Langer N, Hansen LK. Microstate EEGlab toolbox: An introductory guide [Internet]. bioRxiv; 2018. Disponibile su: https://doi.org/10.1101/289850

65. Rabinovici GD. Controversy and Progress in Alzheimer’s Disease - FDA Approval of Aducanumab. N Engl J Med. 26 agosto 2021;385(9):771–4.

66. Shcherbinin S, Evans CD, Lu M, Andersen SW, Pontecorvo MJ, Willis BA, et al. Association of Amyloid Reduction After Donanemab Treatment With Tau Pathology and Clinical Outcomes. JAMA Neurol. ottobre 2022;79(10):1015–24.

67. van Dyck CH, Swanson CJ, Aisen P, Bateman RJ, Chen C, Gee M, et al. Lecanemab in Early Alzheimer’s Disease. N Engl J Med. 29 novembre 2022;

68. Withington CG, Turner RS. Amyloid-Related Imaging Abnormalities With Anti-amyloid Antibodies for the Treatment of Dementia Due to Alzheimer’s Disease. Front Neurol. 23 marzo 2022;13:862369.

69. Clark CM, Pontecorvo MJ, Beach TG, Bedell BJ, Coleman RE, Doraiswamy PM, et al. Cerebral PET with florbetapir compared with neuropathology at autopsy for detection of neuritic amyloid-β plaques: a prospective cohort study. Lancet Neurol. agosto 2012;11(8):669–78.

70. Sabri O, Sabbagh MN, Seibyl J, Barthel H, Akatsu H, Ouchi Y, et al. Florbetaben PET imaging to detect amyloid beta plaques in Alzheimer’s disease: phase 3 study. Alzheimers Dement. agosto 2015;11(8):964–74.

71. Mueller A, Bullich S, Barret O, Madonia J, Berndt M, Papin C, et al. Tau PET imaging with 18F-PI-2620 in Patients with Alzheimer Disease and Healthy Controls: A First-in-Humans Study. J Nucl Med. giugno 2020;61(6):911–9.

72. Lewczuk P, Esselmann H, Otto M, Maler JM, Henkel AW, Henkel MK, et al. Neurochemical diagnosis of Alzheimer’s dementia by CSF Abeta42, Abeta42/Abeta40 ratio and total tau. Neurobiol Aging. marzo 2004;25(3):273–81.

73. Buerger K, Ewers M, Pirttilä T, Zinkowski R, Alafuzoff I, Teipel SJ, et al. CSF phosphorylated tau protein correlates with neocortical neurofibrillary pathology in Alzheimer’s disease. Brain. novembre 2006;129(Pt 11):3035–41.

74. Moschini V, Mazzeo S, Bagnoli S, Padiglioni S, Emiliani F, Giacomucci G, et al. CAG Repeats Within the Non-pathological Range in the HTT Gene Influence Personality Traits in Patients With Subjective Cognitive Decline: A 13-Year Follow-Up Study. Front Psychiatry. 2022;13:826135.

75. Hao L, Sun Y, Li Y, Wang J, Wang Z, Zhang Z, et al. Demographic characteristics and neuropsychological assessments of subjective cognitive decline (SCD) (plus). Ann Clin Transl Neurol. 2020;7(6):1002–12.

76. Wolfsgruber S, Polcher A, Koppara A, Kleineidam L, Frölich L, Peters O, et al. Cerebrospinal Fluid Biomarkers and Clinical Progression in Patients with Subjective Cognitive Decline and Mild Cognitive Impairment. J Alzheimers Dis. 2017;58(3):939–50.

77. Poptsi E, Moraitou D, Tsardoulias E, Symeonidisd AL, Tsolaki M. Is the Discrimination of Subjective Cognitive Decline from Cognitively Healthy Adulthood and Mild Cognitive Impairment Possible? A Pilot Study Utilizing the R4Alz Battery. J Alzheimers Dis. 2020;77(2):715–32.

78. Kurt P, Yener G, Oguz M. Impaired digit span can predict further cognitive decline in older people with subjective memory complaint: a preliminary result. Aging Ment Health. aprile 2011;15(3):364–9.

79. Silva D, Guerreiro M, Maroco J, Santana I, Rodrigues A, Bravo Marques J, et al. Comparison of four verbal memory tests for the diagnosis and predictive value of mild cognitive impairment. Dement Geriatr Cogn Dis Extra. gennaio 2012;2:120–31.

80. Balota DA, Tse CS, Hutchison KA, Spieler DH, Duchek JM, Morris JC. Predicting Conversion to Dementia of the Alzheimer Type in a Healthy Control Sample: The Power of Errors in Stroop Color Naming. Psychol Aging. marzo 2010;25(1):208–18.

81. Van Mierlo LD, Wouters H, Sikkes SAM, Van der Flier WM, Prins ND, Bremer JAE, et al. Screening for Mild Cognitive Impairment and Dementia with Automated, Anonymous Online and Telephone Cognitive Self-Tests. J Alzheimers Dis. 56(1):249–59.

82. Stern Y. Cognitive reserve. Neuropsychologia. agosto 2009;47(10):2015–28.

83. Stern Y. What is cognitive reserve? Theory and research application of the reserve concept. J Int Neuropsychol Soc. marzo 2002;8(3):448–60.

84. Lojo-Seoane C, Facal D, Guàrdia-Olmos J, Juncos-Rabadán O. Structural Model for Estimating the Influence of Cognitive Reserve on Cognitive Performance in Adults with Subjective Memory Complaints. Arch Clin Neuropsychol. 1 maggio 2014;29(3):245–55.

85. Perquin M, Diederich N, Pastore J, Lair ML, Stranges S, Vaillant M, et al. Prevalence of Dementia and Cognitive Complaints in the Context of High Cognitive Reserve: A Population-Based Study. PLOS ONE. set 2015;10(9):e0138818.

86. João AA, Maroco J, Ginó S, Mendes T, Mendonça A de, Martins IP. Education modifies the type of subjective memory complaints in older people. International Journal of Geriatric Psychiatry. 25 maggio 2015;31(2):153–60.

87. Aghjayan SL, Buckley RF, Vannini P, Rentz DM, Jackson JD, Sperling RA, et al. The influence of demographic factors on subjective cognitive concerns and beta-amyloid. International Psychogeriatrics. aprile 2017;29(4):645–52.

88. Lojo-Seoane C, Facal D, Guàrdia-Olmos J, Pereiro AX, Juncos-Rabadán O. Effects of Cognitive Reserve on Cognitive Performance in a Follow-Up Study in Older Adults With Subjective Cognitive Complaints. The Role of Working Memory. Front Aging Neurosci. 2018;10:189.

89. Lee YC, Kang JM, Lee H, Kim K, Kim S, Yu TY, et al. Subjective cognitive decline and subsequent dementia: a nationwide cohort study of 579,710 people aged 66 years in South Korea. Alzheimers Res Ther. 06 2020;12(1):52.

90. Liew TM. Subjective cognitive decline, anxiety symptoms, and the risk of mild cognitive impairment and dementia. Alzheimer’s Research & Therapy. Settembre 2020;12(1):107.

91. Huang Y ping, Xue J jun, Li C, Chen X, Fu H juan, Fei T, et al. Depression and APOEε4 Status in Individuals with Subjective Cognitive Decline: A Meta-Analysis. Psychiatry Investig. 28 agosto 2020;17(9):858–64.

92. Terracciano A, Sutin AR, An Y, O’Brien RJ, Ferrucci L, Zonderman AB, et al. Personality and risk of Alzheimer’s disease: new data and meta-analysis. Alzheimers Dement. marzo 2014;10(2):179–86.

93. Luchetti M, Terracciano A, Stephan Y, Sutin AR. Personality and Cognitive Decline in Older Adults: Data From a Longitudinal Sample and Meta-Analysis. J Gerontol B Psychol Sci Soc Sci. luglio 2016;71(4):591–601.

94. Bessi V, Mazzeo S, Padiglioni S, Piccini C, Nacmias B, Sorbi S, et al. From Subjective Cognitive Decline to Alzheimer’s Disease: The Predictive Role of Neuropsychological Assessment, Personality Traits, and Cognitive Reserve. A 7-Year Follow-Up Study. Journal of Alzheimer’s disease: JAD. 30 maggio 2018;63(4):1523–35.

95. Alberdi A, Aztiria A, Basarab A. On the early diagnosis of Alzheimer’s Disease from multimodal signals: A survey. Artif Intell Med. luglio 2016;71:1–29.

96. Malek N, Baker MR, Mann C, Greene J. Electroencephalographic markers in dementia. Acta Neurol Scand. aprile 2017;135(4):388–93.

97. Gouw AA, Alsema AM, Tijms BM, Borta A, Scheltens P, Stam CJ, et al. EEG spectral analysis as a putative early prognostic biomarker in nondemented, amyloid positive subjects. Neurobiol Aging. settembre 2017;57:133–42.

98. Maestú F, Cuesta P, Hasan O, Fernandéz A, Funke M, Schulz PE. The Importance of the Validation of M/EEG With Current Biomarkers in Alzheimer’s Disease. Frontiers in Human Neuroscience. 2019;13:17.

99. Fonseca LC, Tedrus Gmas, Prandi LR, Andrade ACA de. Quantitative electroencephalography power and coherence measurements in the diagnosis of mild and moderate Alzheimer’s disease. Arq Neuropsiquiatr. 2011;69(2B):297–303.

100. Jelic V, Shigeta M, Julin P, Almkvist O, Winblad B, Wahlund LO. Quantitative electroencephalography power and coherence in Alzheimer’s disease and mild cognitive impairment. Dementia. dicembre 1996;7(6):314–23.

101. Kim JS, Lee SH, Park G, Kim S, Bae SM, Kim DW, et al. Clinical Implications of Quantitative Electroencephalography and Current Source Density in Patients with Alzheimer’s Disease. Brain Topogr. 1 ottobre 2012;25(4):461–74.

102. Kulkarni NileshN, Parhad SaurabhV, Shaikh YasminP. Use of Non-linear and Complexity Features for EEG Based Dementia & Alzheimer Disease Diagnosis. In: 2017 International Conference on Computing, Communication, Control and Automation (ICCUBEA). 2017. p. 1–3.

103. Shumbayawonda E, López-Sanz D, Bruña R, Serrano N, Fernández A, Maestú F, et al. Complexity changes in preclinical Alzheimer’s disease: An MEG study of subjective cognitive decline and mild cognitive impairment. Clinical Neurophysiology. 2020;131(2):437–45.

104. Tait L, Tamagnini F, Stothart G, Barvas E, Monaldini C, Frusciante R, et al. EEG microstate complexity for aiding early diagnosis of Alzheimer’s disease. Scientific Reports. 2020;10(1).

105. Babiloni C, Del Percio C, Lizio R, Noce G, Lopez S, Soricelli A, et al. Functional cortical source connectivity of resting state electroencephalographic alpha rhythms shows similar abnormalities in patients with mild cognitive impairment due to Alzheimer’s and Parkinson’s diseases. Clinical Neurophysiology. 1 aprile 2018;129(4):766–82.

106. Besthorn C, Förstl H, Geiger-Kabisch C, Sattel H, Gasser T, Schreiter-Gasser U. EEG coherence in Alzheimer disease. Electroencephalography and Clinical Neurophysiology. 1 marzo 1994;90(3):242–5.

107. Meghdadi AH, Karić MS, McConnell M, Rupp G, Richard C, Hamilton J, et al. Resting state EEG biomarkers of cognitive decline associated with Alzheimer’s disease and mild cognitive impairment. PLOS ONE. 5 febbraio 2021;16(2):e0244180.

108. Smailovic U, Koenig T, Savitcheva I, Chiotis K, Nordberg A, Blennow K, et al. Regional Disconnection in Alzheimer Dementia and Amyloid-Positive Mild Cognitive Impairment: Association Between EEG Functional Connectivity and Brain Glucose Metabolism. Brain Connect. 1 dicembre 2020;10(10):555–65.

109. Waninger S, Berka C, Meghdadi A, Karic MS, Stevens K, Aguero C, et al. Event-related potentials during sustained attention and memory tasks: Utility as biomarkers for mild cognitive impairment. Alzheimer’s & Dementia: Diagnosis, Assessment & Disease Monitoring. 1 gennaio 2018;10:452–60.

110. Picton TW, Bentin S, Berg P, Donchin E, Hillyard SA, Johnson R, et al. Guidelines for using human event-related potentials to study cognition: recording standards and publication criteria. Psychophysiology. marzo 2000;37(2):127–52.

111. Hillyard SA, Hink RF, Schwent VL, Picton TW. Electrical signs of selective attention in the human brain. Science. 12 ottobre 1973;182(4108):177–80.

112. Polich J, Kok A. Cognitive and biological determinants of P300: an integrative review. Biol Psychol. ottobre 1995;41(2):103–46.

113. Olichney JM, Taylor JR, Gatherwright J, Salmon DP, Bressler AJ, Kutas M, et al. Patients with MCI and N400 or P600 abnormalities are at very high risk for conversion to dementia. Neurology. 6 maggio 2008;70(19 Pt 2):1763–70.

114. Guillaume A, Osmont D, Vartanian A, Gaffie D, Chastres V, Sarron JC, et al. Evaluation of cerebral stresses under acceleration taking into account the lateral ventricles. J Gravit Physiol. luglio 1999;6(1):P67–68.

115. Alexander DM, Arns MW, Paul RH, Rowe DL, Cooper N, Esser AH, et al. Eeg markers for cognitive decline in elderly subjects with subjective memory complaints. J Integr Neurosci. 1 marzo 2006;05(01):49–74.

116. Babiloni C, Visser PJ, Frisoni G, De Deyn PP, Bresciani L, Jelic V, et al. Cortical sources of resting EEG rhythms in mild cognitive impairment and subjective memory complaint. Neurobiology of Aging. 1 ottobre 2010;31(10):1787–98.

117. Jeong HT, Youn YC, Sung HH, Kim SY. Power Spectral Changes of Quantitative EEG in the Subjective Cognitive Decline: Comparison of Community Normal Control Groups. Neuropsychiatr Dis Treat. 24 agosto 2021;17:2783–90.

118. Park JH, Cho HE, Kim JH, Wall MM, Stern Y, Lim H, et al. Machine learning prediction of incidence of Alzheimer’s disease using large-scale administrative health data. npj Digit Med. 26 marzo 2020;3(1):1–7.

119. James C, Ranson JM, Everson R, Llewellyn DJ. Performance of Machine Learning Algorithms for Predicting Progression to Dementia in Memory Clinic Patients. JAMA Network Open. 16 dicembre 2021;4(12):e2136553.

120. Ezzati A, Harvey DJ, Habeck C, Golzar A, Qureshi IA, Zammit AR, et al. Predicting Amyloid-β Levels in Amnestic Mild Cognitive Impairment Using Machine Learning Techniques. J Alzheimers Dis. 2020;73(3):1211–9.

121. Kang SH, Cheon BK, Kim JS, Jang H, Kim HJ, Park KW, et al. Machine Learning for the Prediction of Amyloid Positivity in Amnestic Mild Cognitive Impairment. J Alzheimers Dis. 2021;80(1):143–57.

122. Jo T, Nho K, Saykin AJ. Deep Learning in Alzheimer’s Disease: Diagnostic Classification and Prognostic Prediction Using Neuroimaging Data. Front Aging Neurosci. 2019;11:220.

123. Sarica A, Cerasa A, Quattrone A. Random Forest Algorithm for the Classification of Neuroimaging Data in Alzheimer’s Disease: A Systematic Review. Front Aging Neurosci. 6 ottobre 2017;9:329.

124. Grueso S, Viejo-Sobera R. Machine learning methods for predicting progression from mild cognitive impairment to Alzheimer’s disease dementia: a systematic review. Alzheimer’s Research & Therapy. 28 settembre 2021;13(1):162.

125. Costafreda SG, Dinov ID, Tu Z, Shi Y, Liu CY, Kloszewska I, et al. Automated hippocampal shape analysis predicts the onset of dementia in Mild Cognitive Impairment. Neuroimage. 1 maggio 2011;56(1):212–9.

126. Wee C, Yap P, Shen D. Prediction of Alzheimer’s disease and mild cognitive impairment using cortical morphological patterns. Hum Brain Mapp. 28 agosto 2012;34(12):3411–25.

127. Young J, Modat M, Cardoso MJ, Mendelson A, Cash D, Ourselin S. Accurate multimodal probabilistic prediction of conversion to Alzheimer’s disease in patients with mild cognitive impairment. Neuroimage Clin. 19 maggio 2013;2:735–45.

128. Jack CR, Bennett DA, Blennow K, Carrillo MC, Dunn B, Haeberlein SB, et al. NIA-AA Research Framework: Toward a biological definition of Alzheimer’s disease. Alzheimers Dement. aprile 2018;14(4):535–62.

129. Abdelnour C, Rodríguez-Gómez O, Alegret M, Valero S, Moreno-Grau S, Sanabria Á, et al. Impact of Recruitment Methods in Subjective Cognitive Decline. J Alzheimers Dis. 2017;57(2):625–32.

